# Aeron vs. Gaming Chair - Performance, Perception and Muscle Stiffness in Video Game Players: A Comparative Investigation

**DOI:** 10.1101/2024.03.13.24304245

**Authors:** Joanne DiFrancisco-Donoghue, Hallie Zwibel, William G Werner

**Affiliations:** New York Institute of Technology, College of Osteopathic Medicine (NYITCOM), Department of Osteopathic Medicine, Old Westbury, NY, USA; NYITCOM, Department of Family Medicine, Old Westbury, NY, USA; NYIT School of Health Professions, Department of Physical Therapy, Old Westbury, NY, USA

**Keywords:** gaming, fatigue, back pain, ergonomics, esports

## Abstract

This study compared an Aeron office chair and a commercial gaming chair (GC) on muscle stiffness (MS), performance, and perceptions during a 2-hour gaming session.

Thirty-three esports players (23 ± 4.9) signed consent to participate in this mixed-methods randomized study. Subjects played League of Legends (LoL) in a controlled environment for two 2-hour sessions. MS was measured using oscillation frequency. Investigators recorded evaluations, game statistics, and player perceptions.

Descriptive statistics showed lower MS in the thoracic and lumbar region (left -4.4% vs. 0.32%; -2.7% vs. -2.1%; right 0.2% vs. 8.3%; 7% vs. 10.8%). The upper shoulder was higher in the GC only on the right (9.2% vs. -6.4; left 4.7 vs. 7.5). Most participants preferred the GC (58%), and players won 25% more and achieved 15% more kills in the GC.

The GC exhibited lower levels of muscle stiffness in the thoracic and lumbar regions. This data suggests that the GC is the preferred choice among this group of LoL gamers and is associated with enhanced performance.

## Background

Ergonomic studies that evaluate seats and workstations frequently assess comfort and discomfort. The terms “sitting comfort” and “sitting discomfort” were defined as distinct concepts connected to several aspects: discomfort is related to biomechanics and fatigue concerns, while comfort is connected to a sense of well-being and aesthetics. (1) In other words, a decrease in pain does not necessarily result in a rise in comfort. The absence of discomfort does not necessarily correlate with comfort. Comfort does not always follow from the absence of pain. (2) Generally, it has been theorized that chairs must fit a user’s anthropometrics (body measurements) to eliminate discomfort while sitting. Therefore, gaming chair designs are continuously being modified to reduce discomfort and prevent injuries and disorders, and ergonomic gaming chairs are continually being created. The standard ergonomic recommendation is to try and maintain an upright position to be comfortable sitting in a chair. However, people cannot maintain an erect upright posture for long periods, and fatigue causes discomfort, causing sitters to slouch forward in their seats. (3) The upright posture is only maintained for 15 minutes, according to Fenety et al. (3)

In research conducted by DiFrancisco-Donoghue et al. (4), 42% of collegiate esports players reported experiencing neck and back pain. Additionally, more than 40% of these individuals acknowledged not engaging in any form of physical exercise. A subsequent study by the same team in 2020 revealed that these collegiate esport players had approximately 15% less muscle mass compared to age-matched peers. This convergence of factors heightens the risk of musculoskeletal problems and discomfort during prolonged gaming sessions.

Physical deconditioning, paired with a lack of regular exercise, might predispose these individuals to onset neck pain (NP) stemming from poor postural habits. A leading cause of chronic neck pain (CNP) is the forward protraction of the head, resulting in accentuated lordosis in the cervical region and subsequent muscle weakening. This positioning can further lead to complications like neuromuscular dysfunction, as noted by Kang & Kim.(5) Such impairments could detrimentally affect esports players by disrupting focus, slowing physical response time due to neuromuscular dysfunction, and reducing their endurance and capacity to play for extended durations.

While examining office chairs, the degree of discomfort rose over time sitting and was unrelated to chair design.(2) Previous studies have found a higher rating of comfort in chairs that people felt were appealing in style and well-made and noted two identical chairs would elicit different ratings of comfort, depending on the aesthetics of the cloth material used to cover the chairs.(1) There is no literature, however, that has examined any of these factors on comfort and discomfort in gaming chairs and how this may impact performance.

MS and discomfort while gaming for prolonged periods is best assessed in the natural gaming environment because it is dependent on the task of the game as well as a variety of other factors related to the individual personal setup preferences. (2)

This study investigates how chair design differences between a typical Aeron office chair and a specific gaming chair influence discomfort during extended gaming sessions for 2 hours. Additionally, it assesses the impact of chair design on muscular postural characteristics, particularly MS. Other objectives are to evaluate gaming performance based on chair type and ascertain whether the chair design affects subjective feelings of comfort.

## METHODS

All subjects were recruited between November 21, 2022 through April 25, 2023. Thirty-three healthy competitive esport players (age 23 ± 4.9) signed written informed consent to participate in this mixed-methods randomized trial. The New York Institute of Technology (NYIT) Institutional Review Board approved the study. *Inclusion Criteria:* A ranked esport player in League of Legends over the age of 18. Exclusion criteria: any known history of a musculoskeletal injury to the back or upper body.

### Procedures

This study was a mixed-method randomized cross-over design trial that required subjects to come to the esport gaming lab for two testing days lasting 2.5 hours. The room was temperature controlled for All subjects and kept within 2-3 degrees Celsius each testing day. Before each gaming session, subjects were fitted to each chair according to manufacturer guidelines. Participants were also shown each chair’s adjustable features (e.g., height adjustment, lumbar adjustment, arm adjustment, and head support). The subjects were given the exact instructions each day: “Please adjust the chair to your preference before gaming.”

#### Testing day 1

Following consent and pre-surveys, all subjects played continuously for 2 hours on day one with no break. Players were fitted to the proper seat chair size as per manufacturer guidelines. Each subject was asked to adjust the chair to their preference of height, back adjustment, and distance to the monitor. Soft tissue changes on the upper trapezius muscle, the mid trapezius muscle, the lower trapezius muscle, and the lower erector spinae muscle were tested before gameplay. (see Figure 2). All samples were bi-lateral pre-play and at 2 hours. After 2 hours of game, the subjects completed the Chair Evaluation Checklist, open-ended survey, and perception questions. Wins, losses, kills, and scores were recorded for each game. On each testing day, all subjects were asked to play against the same level and game rank.

#### Day 2

All methods were repeated with the other chair on another day with at least two days’ rest.

## Outcomes

### Muscle Stiffness (MS)

MS was measured using the MyotonPRO™ device (Myoton AS, Tallinn, Estonia). The MyotonPRO™ is a portable device for measuring biomechanical and viscoelastic properties in superficial soft tissues. This device uses oscillation frequency (Hz) to give a quantitative value of viscoelastic properties. As a muscle increases intramuscular pressure by being in a constant state of tension, it has reduced blood supply and fatigue. This device uses silent EMG signals to detect these changes. (6,7) This device has been validated in assessing wrist stiffness and upper and lower body muscles. See **Figure 1** anatomical sites.

**Figure 1.**
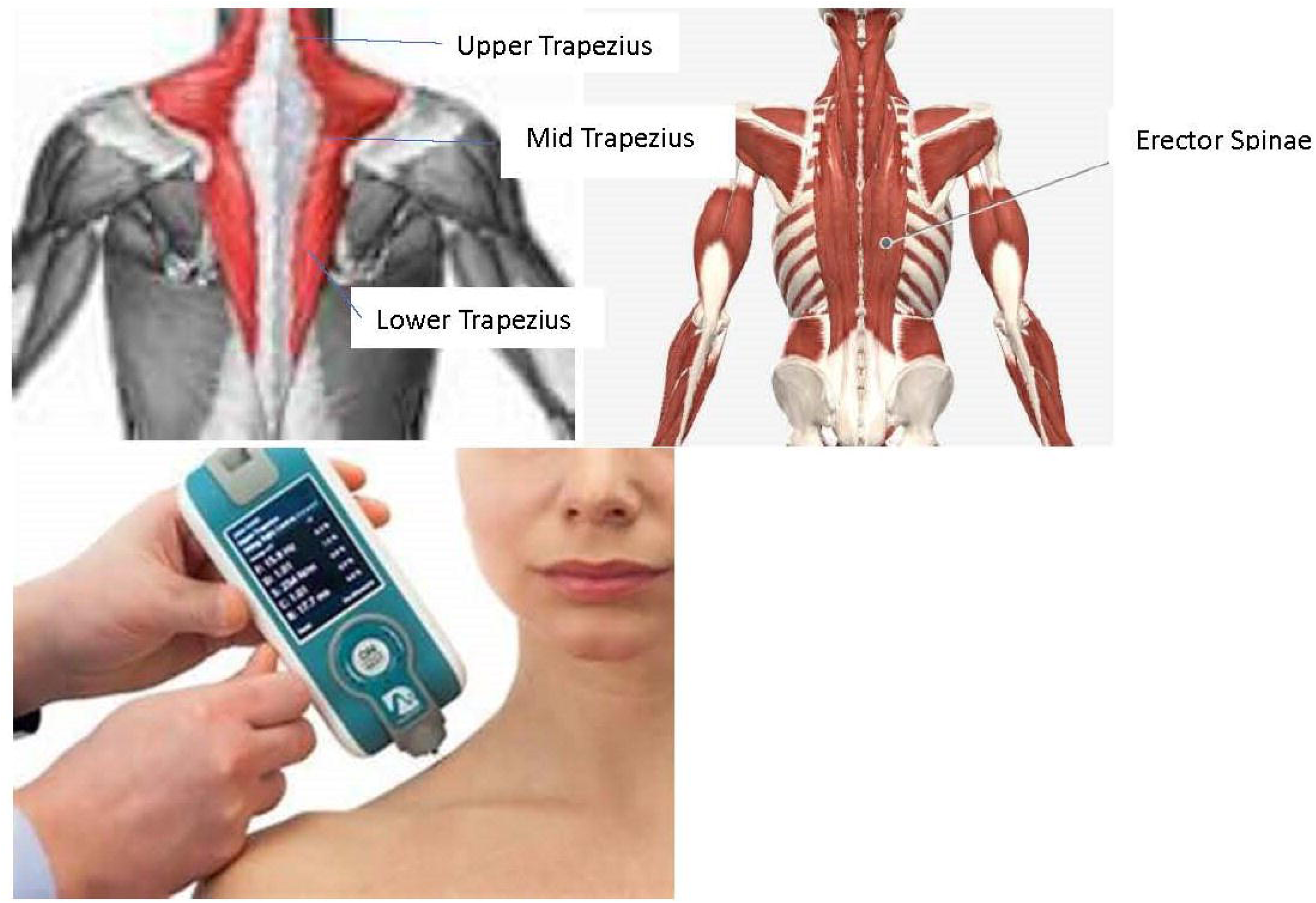
Myoton™ device with anatomical testing sites done bilaterally.

**Figure 2.**
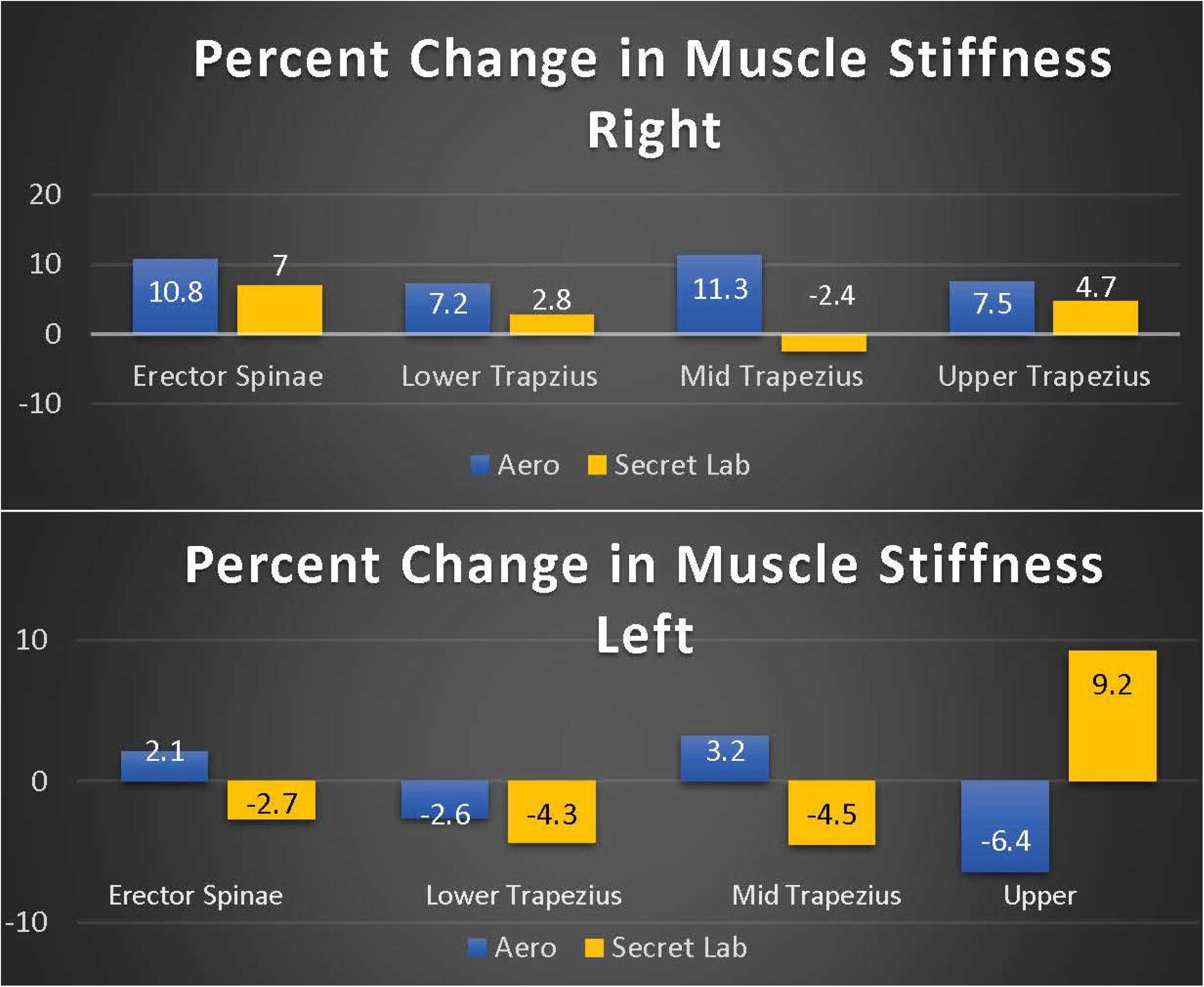
Percent difference in MS by side following 2 hours of LoL gameplay.

### The Chair Checklist Survey(CCS)

The version of the CCS used in this study was adopted by Helander et al.(1). It consists of 13 descriptors of comfort and discomfort.

### Open-ended Subject Perception Survey

Following each gaming session, a survey was conducted to learn how the respondents perceived each chair. A final survey was conducted to understand how the subjects perceived both chairs.

#### Statistical Analysis

This study used a repeated measure followed by a post hoc analysis to compare MS on the four bilateral testing sites between the two chair conditions with a significance set at 0.05. A non-parametric analysis, Wilcoxon signed-rank test, was used to compare the two chair conditions for the Cornell Musculoskeletal Discomfort Questionnaire (CMDQ) and the Chair Evaluation Checklist (CEC). For the General Comfort Rating Scale (GCRS), a McNemar test was used to compare the chairs. The open-ended survey used Descriptive statistics between the two testing chair groups.

## Results

No significant effects were found between MS in the erector spinae, lower trapezius, and mid trapezius on the left side. A difference was found in the upper trapezius between groups on the left side (p= 0.03). There were no significant differences found between groups on the right side for all four muscles. Descriptive statistics revealed that the right-side gaming chair had 3.8% less MS in the erector spinae muscle, 4.4% less MS in the lower trapezius, 13.7% less MS in the mid-trapezius muscle, and 2.8% less MS in the upper trapezius. On the left side, the gaming chair revealed 4.8% less MS, 1.7% less in the lower trapezius, 7.7% less in the mid trapezius, and a 15.6 % increase in MS in the upper trapezius.

The chair checklist survey revealed a significant difference in “The chair looks nice” between chair groups, with 29% of participants favoring the gaming chair, p= 0.003, 18% of participants felt more relaxed in the gaming chair, p=0.037; and 20% of participants felt the gaming chair was more spacious than the Aeron chair, p=0.018. There were no other significant differences found in the Chair Checklist Survey; full descriptive and statical results are presented in **Table 3**.

**Table 1.**
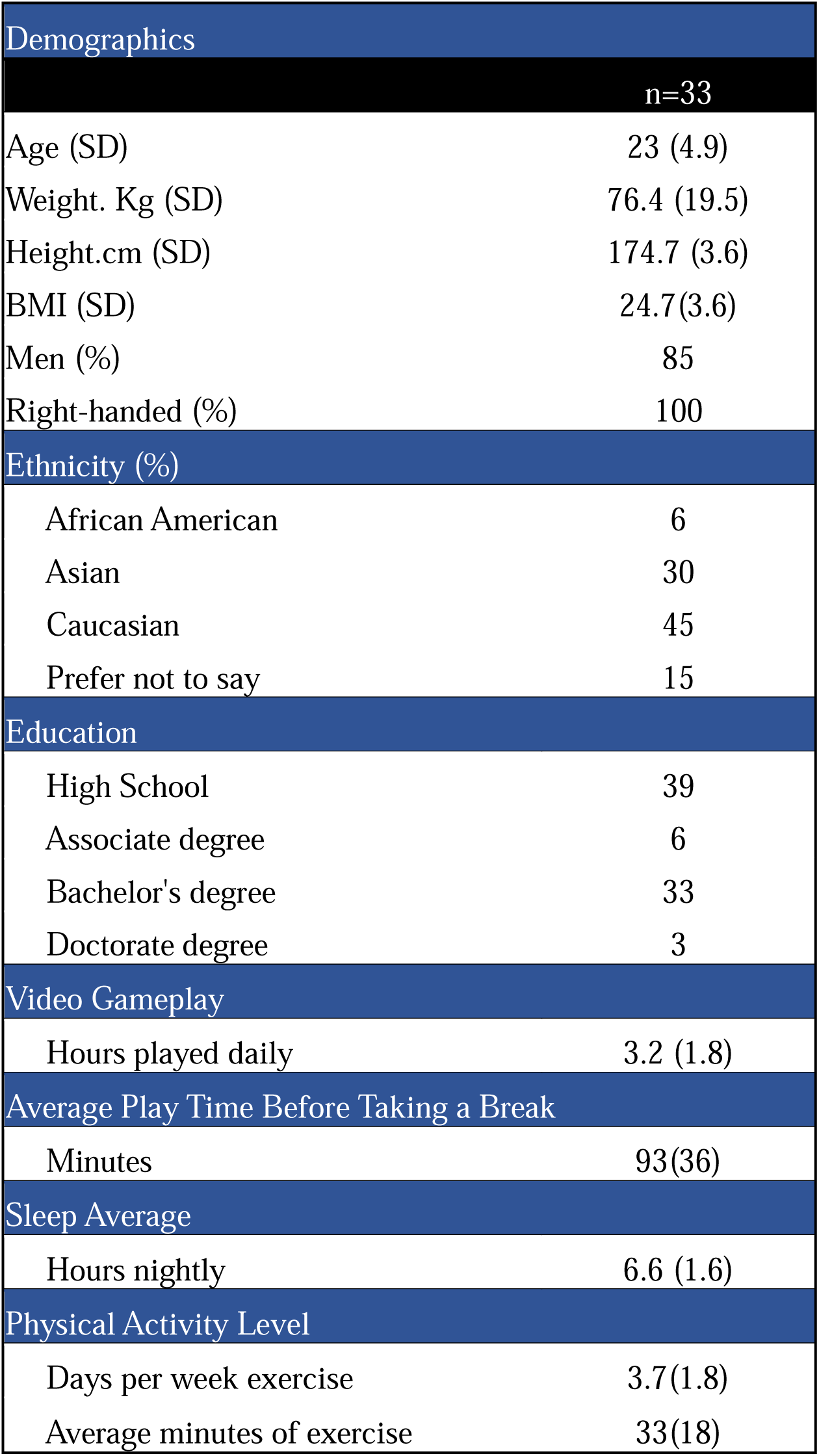
Subject Demographics.

**Table 2.**
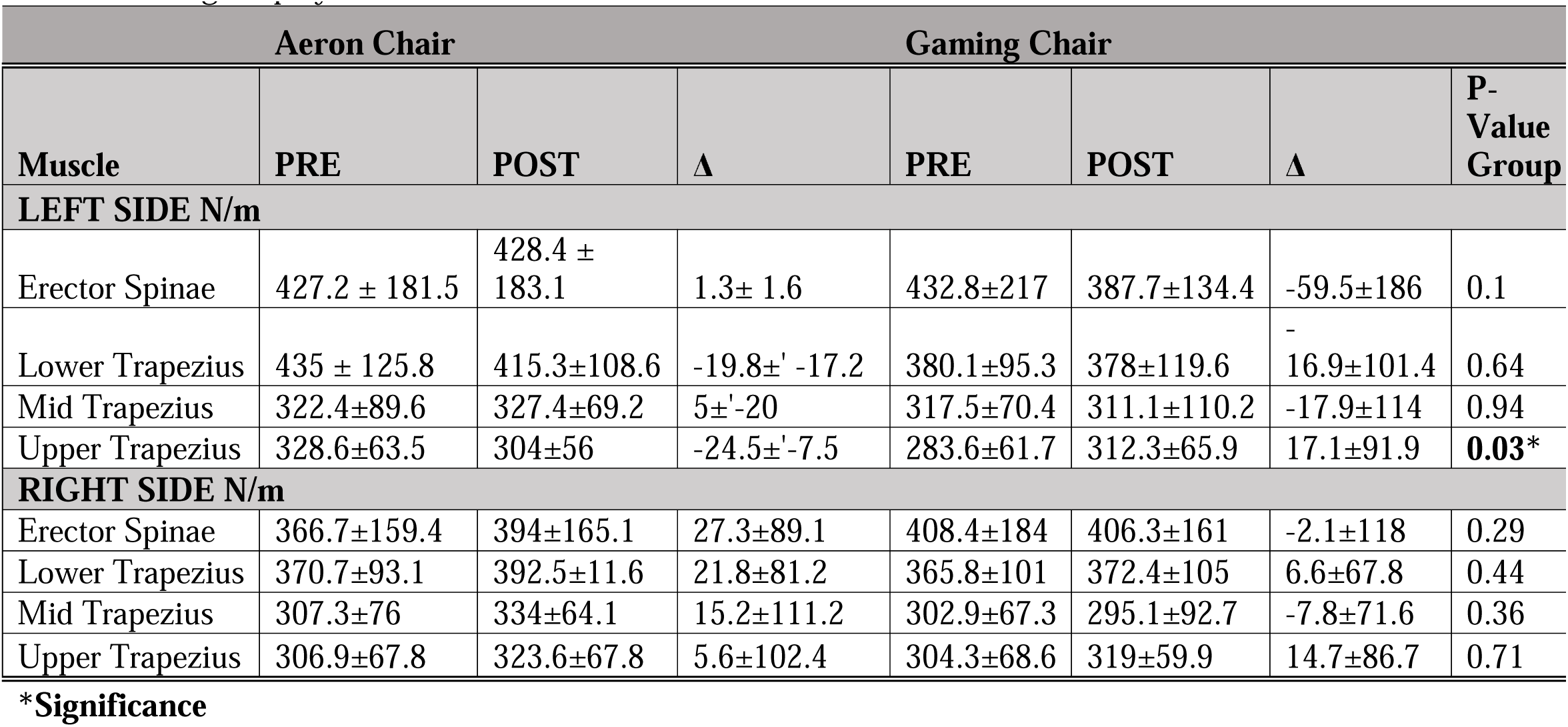
Muscle stiffness pre and post-by-side following 2 of LoL gameplay.

**Table 3.** Results of the Chair Checklist Survey (CCS)

| CCS Item | Aeron | Gaming Chair (GC) | p-value | % Difference |
| --- | --- | --- | --- | --- |
| I have sore muscles | 2.7 (1.8) | 2.2 (1.3) | 0.27 | 20% less sore muscles in GC |
| **The chair looks nice** | **5.3 (2.1)** | **7.1 (1.5)** | **0.003\*** | **29% more favored GC** |
| I have heavy legs | 3.0 (2.3) | 2.2 (1.6) | 0.16 | 31% less heavy legs in GC |
| I feel stiff | 3.1 (2.0) | 2.9 (1.5) | 0.97 | 6.7% less stiff in GC |
| I like the chair | 6.0 (2.1) | 7.0 (1.7) | 0.05 | 15% liked GC more |
| I feel restless | 2.9 (2.3) | 3.1 (2.3) | 0.75 | 7% more restless in GC |
| The chair feels soft | 5.0 (2.2) | 5.3 (2.3) | 0.62 | 6% more soft GC |
| I feel tired | 2.4 (2.0) | 2.2 (1.6) | 0.50 | 9% less tired in GC |
| I feel pain | 2.3 (1.9) | 1.8 (1.1) | 0.26 | 24% less pain in GC |
| **I feel relaxed** | **5.1 (1.6)** | **6.1 (1.6)** | **0.037\*** | **18% more relaxed in GC** |
| I feel numb | 1.6 (1.4) | 1.5 (1.0) | 0.88 | 7% less numbness in GC |
| I feel uneven pressure | 2.8 (2.1) | 2.4 (1.5) | 0.52 | 15% felt less uneven pressure in GC |
| **The chair is spacious** | **6.0 (1.7)** | **7.3 (1.8)** | **0.018\*** | **20% felt more spacious in GC** |
| I feel cramped | 1.9 (1.8) | 1.6 (0.9) | 0.53 | 17% felt less cramped in GC |
| I feel refreshed | 4.1 (2.3) | 4.9 (1.7) | 0.19 | 18% felt more refreshed in GC |
| I feel restful | 3.7 (2.1) | 4.9 (1.8) | 0.05 | 28% felt more restful in GC |
| I feel comfortable | 6.1 (2.1) | 6.9 (1.5) | 0.12 | 12% felt more comfortable in GC |

Players demonstrated 25% more wins in the gaming chair than the Aeron chair and 15% more kills in the gaming chair compared to the Aeron chair. **Figure 3**.

**Figure 3.**
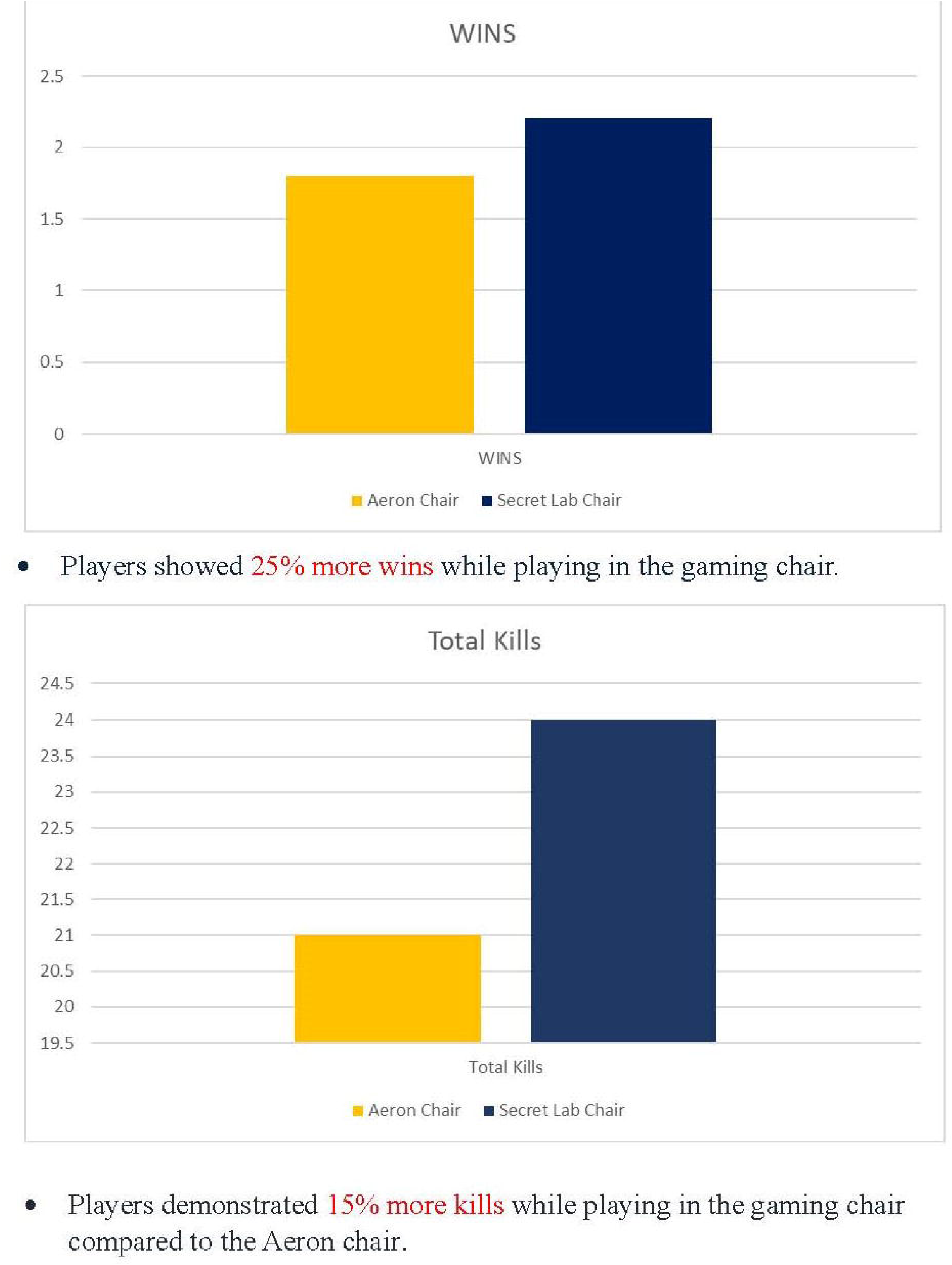
Player Performance by Chair

The results of the look and preference of each chair can be found in **Table 4**.

**Table 4.**
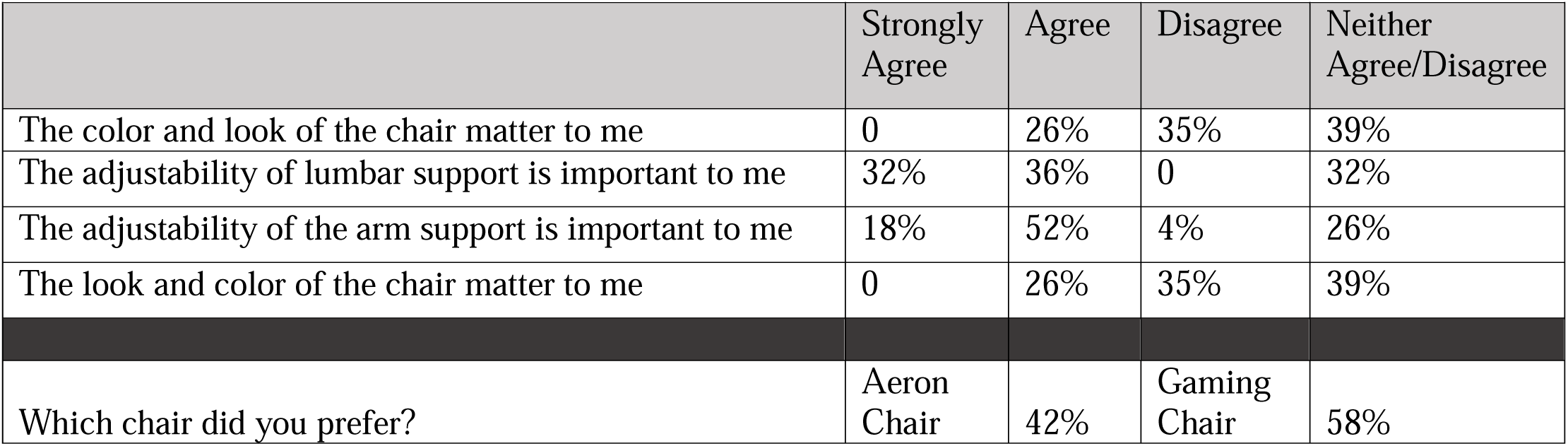
Subject preferences.

Descriptive data on which parts of the chair were enjoyed most and which were enjoyed least can be found in **Figure 4**.

**Figure 4.**
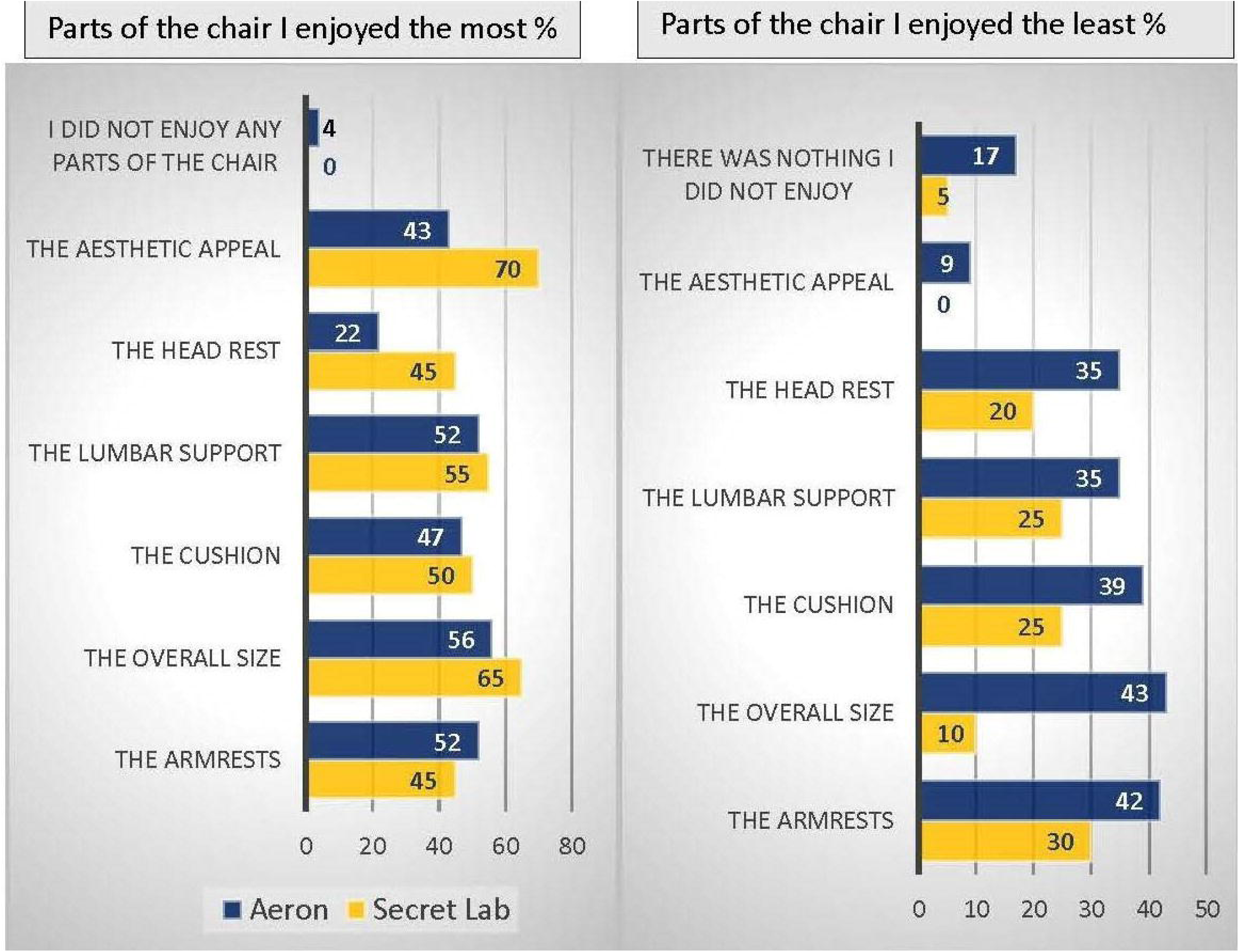
Percentage of participants’ perceptions of what was enjoyed most about each chair and what was least enjoyed.

## Discussion

The impact of ergonomic seating on performance, comfort, and muscle function has gained traction in recent years, particularly with the popularity of esports. This study specifically sought to discern differences in MS, overall comfort, and gaming performance when using a conventional office Aeron chair compared to specialized gaming chairs.

One of the study’s primary findings was the change in MS between chairs. The left upper trapezius is a muscle heavily involved in supporting the dominant arm during repetitive movements, such as mouse use or key pressing. The absence of significant findings in the right upper trapezius suggests a potential interaction between chair ergonomics and handedness that warrants further exploration.

The gaming chair overall in the mid-thoracic and lumbar region showed lower muscle stiffness. With 4.1% less in the lumbar spine and 13.8% less in the thoracic spine after a prolonged sitting period of 2 h. In the context of gaming activity, it’s worth noting that the trapezius muscles play a significant role in the movement and stability of the scapula. Specifically, the lower trapezius muscle assumes a vital role in stabilizing the scapula.(8,9) Prolonged hyperactivation and shortening of this muscle can increase stiffness. The reduced muscle stiffness, particularly in the lower trapezius muscle, suggests that the gaming chair may provide better comfort and support for the spine’s mid-thoracic and lumbar regions during extended gaming over the Aeron chair.

It’s worth noting that subjective feedback from participants revealed a preference for the aesthetic design of the gaming chair. The connection between a chair’s aesthetic appeal and its perceived comfort, as suggested by Helander & Zhang,(1) was echoed in our findings. This phenomenon implies a psychological facet to comfort that transcends biomechanical considerations. It reinforces the idea that perceptions of comfort and well-being are not solely determined by objective physiological metrics but also by personal and cultural aesthetic preferences.

Interestingly, players recorded better game outcomes when using the gaming chair, with 25% more wins and 15% more kills than when using the Aeron chair. Though the direct cause of this improved performance is not wholly evident from this study, it could be inferred that the psychological comfort, alongside potential biomechanical advantages of the gaming chair, combined to create an environment where players could perform at their peak. This potential synergy between aesthetics, physical comfort, and performance is an intriguing area for future research.

However, the study had a few limitations. The absence of a broader range of physical metrics— like posture monitoring, detailed fatigue markers, or even real-time muscle activity recordings— might have provided additional insights. Moreover, the choice of game (League of Legends) may induce specific biomechanical demands that might not generalize to other esports games.

Studies conducted on workspace modifications have shown that the chair directly influences body alignment and posture.(10,11) However, individuals who experience musculoskeletal pain or discomfort are often advised to make frequent adjustments to their workstation chairs.(1,11) Modifying a chair is a practical and viable step to help alleviate gaming discomfort. Therefore, when selecting a chair individuals should consider adjustability of seat height, and seat pan depth that align with their anthropometric and lumbar support adjustments along with armrest adjustments.(11) Chairs that offer these options may play a significant role in preventing discomfort during prolonged gaming.

The gaming chair used in this study offered adjustable arms (both lateral and vertical) with different surface options, as well as adjustments for lumbar support, headrest, height adjustment, and reclining options. This gaming chair was also offered in two sizes based on height and weight. The Aeron chair was one size, offering height and lumbar adjustment with no headrest or reclining option. The armrests are adjusted only vertically. Based on the data collected, the gaming chair is more preferred and comfortable than the Aeron chair for LoL players. Furthermore, most players found the adjustability of the lumbar support and armrests to be important when selecting a chair. Overall, as gaming continues its rapid growth in popularity, it becomes increasingly vital to explore how chair ergonomics impacts musculoskeletal health and overall gaming performance.

This study was funded by Secret Lab™ Inc. under grant #589267

## Data Availability

Data are available by request for researchers who meet the criteria for access to confidential data.

